# Acute renal injury in patients with COVID-19, in the critical care unit of a public hospital, Lima-Peru

**DOI:** 10.1101/2023.08.20.23294334

**Authors:** Yanissa Venegas-Justiniano, Abdías Hurtado-Aréstegui, Karina Mucho-Vilca

## Abstract

**Objective:** To determine the clinical and laboratory characteristics, as well as evaluating the factors associated with mortality in patients with COVID-19 infection and acute kidney injury (AKI) hospitalized in the Intensive Care Unit (ICU) of the Hospital Nacional Arzobispo Loayza.

**Materials and Methods:** Retrospective cohort study, with convenience sampling during the period from April 2020 to March 2021, through the review of medical records data. Inclusion criteria were; patients ≥ 18 years old, with a diagnosis of COVID-19 infection, who were admitted to ICU with normal renal function and developed AKI during their stay in ICU. Exclusion criteria were; patients who developed AKI prior to ICU admission, patients with chronic kidney disease with and without dialysis.

**Results:** A total of 177 medical records that met the inclusion and exclusion criteria were evaluated. The mean age was 57.2±13.2 years, 145 (81.4%) were male; comorbidities were: obesity 112(63.3%), arterial hypertension 55 (31.1%) and diabetes mellitus 30(16.9%); the most frequent cause of AKI was hypoperfusion (93%). 83 participants (46.8%) received dialytic support in the intermittent hemodialysis modality. In-hospital mortality was 151 (85.3%) and was higher in the group with stage 3 AKI: 109 (72.2%). The increase in ferritin level (OR: 10.04 (95%CI 4.4-38.46), p<0.001) and APACHE score (OR: 1.75 (95%CI 1.4-2.12), p<0.001), as well as the decrease in PaO_2_/FiO_2_ level (OR: 0.85 (95%CI 0.59-0.92), p<0.041, were related to mortality.

**Conclusions:** AKI in ICU patients with COVID-19 infection has a high mortality and the related factors were the increase in APACHE II score and ferritin level, as well as the decrease in PaO_2_/FiO_2_ level.

## INTRODUCTION

Since the first case of COVID-19 infection was reported, millions of people have been affected, and it has been associated with high morbidity and mortality. The clinical presentation can vary in intensity, from asymptomatic cases to severe cases with acute respiratory distress syndrome and involvement of other organs and systems (cardiovascular, digestive, renal, hematological and nervous), requiring hospitalization, intensive care unit (ICU) treatment, mechanical ventilation and hemodynamic support (1-6).

The incidence of acute kidney injury (AKI) in COVID-19 is varied and depends on the population evaluated. AKI is more frequent in critically ill patients with COVID-19, affecting between 20-40% of those admitted to the ICU, it is considered a marker of disease severity and lower survival (5-9); it is also associated with a hospital mortality of 45% compared to 7% among those who did not have AKI, requiring renal replacement therapy between 9-14.3% (4,9-14).

Renal injury by COVID 19 has multiple causes and presentations. It can be related to: direct virus aggression (tubular damage and glomerular injury), hypoperfusion, excessive inflammatory response, hypoxia, mechanical ventilation, endothelial damage with microthrombi and nephrotoxicity. The histological findings observed are: tubular necrosis, endothelial damage, capillary erythroid aggregates, glomerular intracapillary fibrin thrombi and inflammatory signs (12,15-18).

Risk factors for AKI include: advanced age, diabetes mellitus, cardiovascular disease, black race, hypertension, need for ventilation, use of vasopressor drugs, and inflammatory markers (18).

The main objective of the study was to describe the clinical laboratory characteristics and factors associated with mortality in patients with COVID-19 infection who presented AKI during hospitalization in the ICU of a public hospital in Lima, Peru.

## MATERIALS AND METHODS

The present study is a retrospective cohort. The study population was selected by non-probabilistic sampling; all cases of COVID-19 in adults admitted to the ICU of the Hospital Nacional Arzobispo Loayza (HNAL) who presented deterioration of renal function between April 1, 2020 and March 31, 2021 were evaluated. HNAL, located in Lima-Peru was one of the main national referral hospitals during the pandemic. The diagnosis of COVID-19 was based on clinical and radiological criteria established by the World Health Organization and was confirmed by detection of SARS-CoV-2 in nasopharyngeal exudate by reverse transcriptase-polymerase chain reaction (RT-PCR) or antigenic testing. No vaccine for COVID-19 was available during the period described.

Inclusion criteria were: patients hospitalized in ICU older than 18 years, with diagnosis of COVID-19 infection, who developed AKI during the hospitalization period. Exclusion criteria were: patients with a diagnosis of chronic kidney disease (any stage), patients with chronic dialysis support and post-transplanted patients.

During the study period, 500 patients with a diagnosis of COVID-19 infection were admitted to the ICU; 184 (36.8%) patients with this diagnosis developed AKI during hospitalization, of whom, 177 met the inclusion criteria. The information was recorded retrospectively, and the clinical records were the primary source of information.

Patients who required admission to the ICU were admitted for severe pulmonary involvement (severe pneumonia and acute respiratory distress syndrome). Mechanical ventilation included invasive and non-invasive methods. The following laboratory variables were considered: leukocytes, lymphocytes, platelets, D-dimer, ferritin, C-reactive protein (CRP), lactate dehydrogenase (LDH), lactate, pH, bicarbonate, potassium, aspartate aminotransferase (AST), alanine aminotransferase (ALT) and arterial oxygen pressure / inspired oxygen fraction (PaO_2_/FiO_2_). Normal values were hospital laboratory reference values and were recorded within the first 48 hours of ICU admission. Severity scores were included: Acute Physiology and Chronic Health Evaluation (APACHE II) and Sequential Organ Failure Assessment score (SOFA). AKI was classified into 3 stages according to the KDIGO guidelines (19), taking the highest serum creatinine value recorded during hospitalization. Renal recovery was considered when serum creatinine returned to the initial or baseline value or lower. Non-recovery was established when creatinine did not decrease or the patient remained on dialysis. The renal support therapy used was intermittent hemodialysis. The variable outcome was death.

### Statistical análisis

Clinical characteristics, inflammation markers, severity score, as well as the clinical outcome of patients with COVID-19 infection and AKI in the ICU were described. The measures of central tendency used were: mean plus standard deviation (SD) for variables with normal distribution, as well as median and interquartile range (IQR) for those without normal distribution. Normality was assessed with the histogram graph and the Shapiro-Wilks test. The Chi-square test was used to compare categorical variables or proportions. To compare two means of independent samples with normal distribution, the Student’s t test was used, and when they did not have normal distribution, the Mann-Whitney U test was used. For the comparison of more than two means, variance analysis (ANOVA) and Kruskal Wallis tests were used depending on whether the data had normal or non-normal distribution, respectively. Logistic regression analysis was used to evaluate the variables related to mortality, with compliance with the following assumptions: dichotomous outcome, independence between observations and linear relationship between the logarithm of the odds and the covariates. In relation to missing data, they represented < 4% of the sample. Data processing and statistical analysis was performed with Stata Software Version 17, with p <0.05 considered statistically significant.

### Ethical Aspects

A review of the medical records and the electronic laboratory database of the Hospital Nacional Arzobispo Loayza was carried out; no direct intervention was made on the patient, so there was no damage derived from the research. The data collection instrument did not include the patient’s name. The protocol was approved by the Ethics Committee of the Hospital Nacional Arzobispo Loayza.

## RESULTS

During the study period, 177 patients with a diagnosis of COVID-19 infection who developed AKI were admitted to the ICU, 72% were stage 3, mean age 57.2±13.2 years, 81% male; the most frequent cause of AKI was hypoperfusion in 93%. In the group with stage 3 AKI there was more: oliguria (80.2% vs 15.6%, p=0.023), need for dialysis support (96.8% vs 3.6%, <0.001) and CRP level (16.4 mg/dl vs 11.5 mg/dl, p<0.041), as well as less time between admission and AKI presentation (9 days vs 11 days, p=0.017). Hemodialysis was in the intermittent modality and was required by 46.9% of patients **(Table 1)**, the start of dialysis therapy was at 13.6 days RIQ [11.3-15.7] and the median duration of therapy was 4.3 days RIQ [3-5]. All received mechanical ventilation and were subjected to prone position.

**Table 1:** Clinical and laboratory characteristics in patients with COVID-19 and AKI in ICU. †.

| Characteristic | n (177) | AKI stage 1<br>n=9 ( 5%) | AKI stage 2<br>n=41(23%) | AKI stage 3<br>n=127(72%) | p |
| --- | --- | --- | --- | --- | --- |
| Male* | 144 (81.4%) | 6(4.2%) | 35(24.3%) | 107(71.5%) | 0.393 |
| Age (years)** | 57.2±13.2 | 57.8±8.4 | 55.1±13.2 | 57.8±13.5 | 0.491 |
| Body mass index *** | 31.2[29-34.3] | 32.9[27.5-36.7] | 31.1[29.1-32.3] | 31.2[28.8-34.6] | 0.442 |
| **Comorbidities\*** |  |  |  |  |  |
| Obesity | 133 (63.3%) | 4(3.1%) | 33(25.8%) | 96(73.3%) | 0.511 |
| Hypertension | 54(31.1%) | 3(3.4%) | 17(32.1%) | 34(62.5%) |  |
| Diabetes mellitus | 30(16.9%) | 2(6.7%) | 10(23.3%) | 18(60%) |  |
| **Hospitalization** |  |  |  |  |  |
| Oliguria * | 96(54.2%) | 4(4.2%) | 15(15.6%) | 77(80.2%) | 0.023 |
| Vasoactive * | 125(70.6%) | 7(5.6%) | 28(22.4%) | 90(72%) | 0.915 |
| Hemodialysis * | 83(46.9%) | 0 | 3(3.6%) | 61(96.8%) | <0.001 |
| APACHE (a)*** | 18 [16-19] | 18[17-21] | 18[16-19] | 18[16-19] | 0.581 |
| SOFA (b)*** | 10[8-11] | 10[8-10] | 10[8-11] | 10[8-11] | 0.933 |
| PEEP(c)*** | 13[12-14] | 12[12-13] | 13[12-14] | 12[12-14] | 0.719 |
| ICU stay (days)*** | 20 [13-28] | 33 [19-46] | 21[14-26] | 19[12-28] | 0.126 |
| Recovery of renal function * | 30 (16.9%) | 1 (33.3%) | 6(20%) | 23(76.7%) | 0.781 |
| Deceased * | 151 (85.3%) | 8(5.3%) | 34(22.5%) | 109(72.2%) | 0.859 |
| Time between admission and presentation of AKI (days)*** | 10 [5-18] | 15 [13-37] | 11 [6-19] | 9[4-16] | 0.017 |
| **Laboratory** |  |  |  |  |  |
| Creatinine at admission mg/dl *** | 0.8 [0.7-0.9] | 0.71[0.6-0.7] | 0.8 [0.7-0.9] | 0.85[0.8-0.9] | 0.004 |
| Creatinine at diagnosis of AKI mg/dl *** | 3.0[2.2-4.5] | 1.4[1.3-1.5] | 2.1[1.8-2.5] | 3.9 [2.8-5.1] | <0.001 |
| Urea mg/dl*** | 40.2[31.3-59.4] | 36.2[25-36.2] | 32.7[27.2-43.2] | 43.5[33.8-69] | <0.001 |
| Leukocytes cells x mm<sup>3</sup>*** | 14400[9630-19692] | 14770[4410-24000] | 12300[9630-17420] | 14600[9690-20000] | 0.060 |
| Lymphocytes cells x mm<sup>3</sup>*** | 860[600-1170] | 960[520-1050] | 920[720-1060] | 530[400-1300] | 0.602 |
| Platelets cells x mm<sup>3</sup>*** | 280000[201000-359000] | 333000[201000-333000] | 280000[201000-335000] | 280000[203000-365000] | 0.852 |
| D-dimer mg/L***† | 1.4[0.5-5.1] | 0.6[0.6-2.2] | 1.6[0.4-3.5] | 1.9[0.6-5.1] | 0.252 |
| Ferritin ng/ml*** | 1578[1000-2000] | 1534[1053-1900] | 1203[989-1901] | 1664[1100-2000] | 0.256 |
| CRP mg/dl (d)*** | 16[9.3-24.3] | 8.7[5.1-23.4] | 10.2[7.5-24.3] | 16.4[10.1-24.1] | 0.041 |
| LDH U/L (e)*** | 584[473-651] | 533[350-712] | 632[473-727] | 601[473-699] | 0.156 |
| Lactate mmol/L*** | 1.8 [1.5-2.3] | 1.7[1.6-1.8] | 1.9[1.6-2.3] | 1.9[1.5-2.4] | 0.439 |
| pH*** | 7.2[7.1-7.3] | 7.1[7.1-7.3] | 7.2[7.1-7.3] | 7.2[7.2-7.3] | 0.911 |
| Bicarbonate mmol/L*** | 19.3[17.9-21.4] | 17.5[15-19.4] | 20[18.5-21.4] | 19[17.9-21.4] | 0.079 |
| Potassium mmol/L*** | 4.1[3.6-4.3] | 3.9[3.6-4.2] | 4.2[3.6-4.1] | 4.1[3.6-4.3] | 0.541 |
| AST U/L (f)***† | 58[35.9-76] | 62[60-71] | 62[40-86] | 50[35-72] | 0.343 |
| ALT U/L (g)***† | 61[40-89] | 78[72-91] | 63[38-109] | 55[40-87] | 0.242 |
| PaO<sub>2</sub>/FiO<sub>2</sub> mmHg (h)*** | 113[98-138] | 126[85-131] | 119[90-150] | 112[98-132] | 0.896 |

(a): Scale of prediction: Acute Physiology And Chronic Health Evaluation; (b): Sequential Organ Failure Assessment score, (e): Positive end-expiratory pressure, (d): C-reactive protein, (e): Lactate dehydrogenase, (f): aspartate aminotransferase, (g): alanine aminotransferase, (h): Arterial Oxygen Pressure / Inspired Oxygen Fraction.
\*: Fisher's exact, \*\*: ANOVA, \*\*\*: Kruskal Wallis
†: Some variables may add up to less than 177 due to missing data.

In-hospital mortality in the study group was 151 (85.3%). Bivariate analysis of living vs deceased patients showed that oliguria (38.5% vs 56.9%, p<0.001), vasoactive use (38.5% vs 76.2%, p<0. 001), Body mass index (29.1 Kg/m^2^ vs 31.5 Kg/m^2^, p=0.019, APACHE score (11 vs 18, p<0.001), SOFA score (7 vs 10, p<0.001), ferritin (867 ng/dl vs 1684 ng/dl, p<0. 001), LDH (400 U/L vs 588 U/L, p<0.001), lactate (1.2 vs 1.9, p<0.001) were higher in the deceased, while pH (7.3 vs 7.2, p<0.001), HCO_3_ (20.7 mmol/L vs 19 mmol/L, p=0.017) and PaO_2_/FiO_2_ (178 mmHg vs 110 mmHg, p<0.001), were lower **(Table 2)**.

**Table 2:**
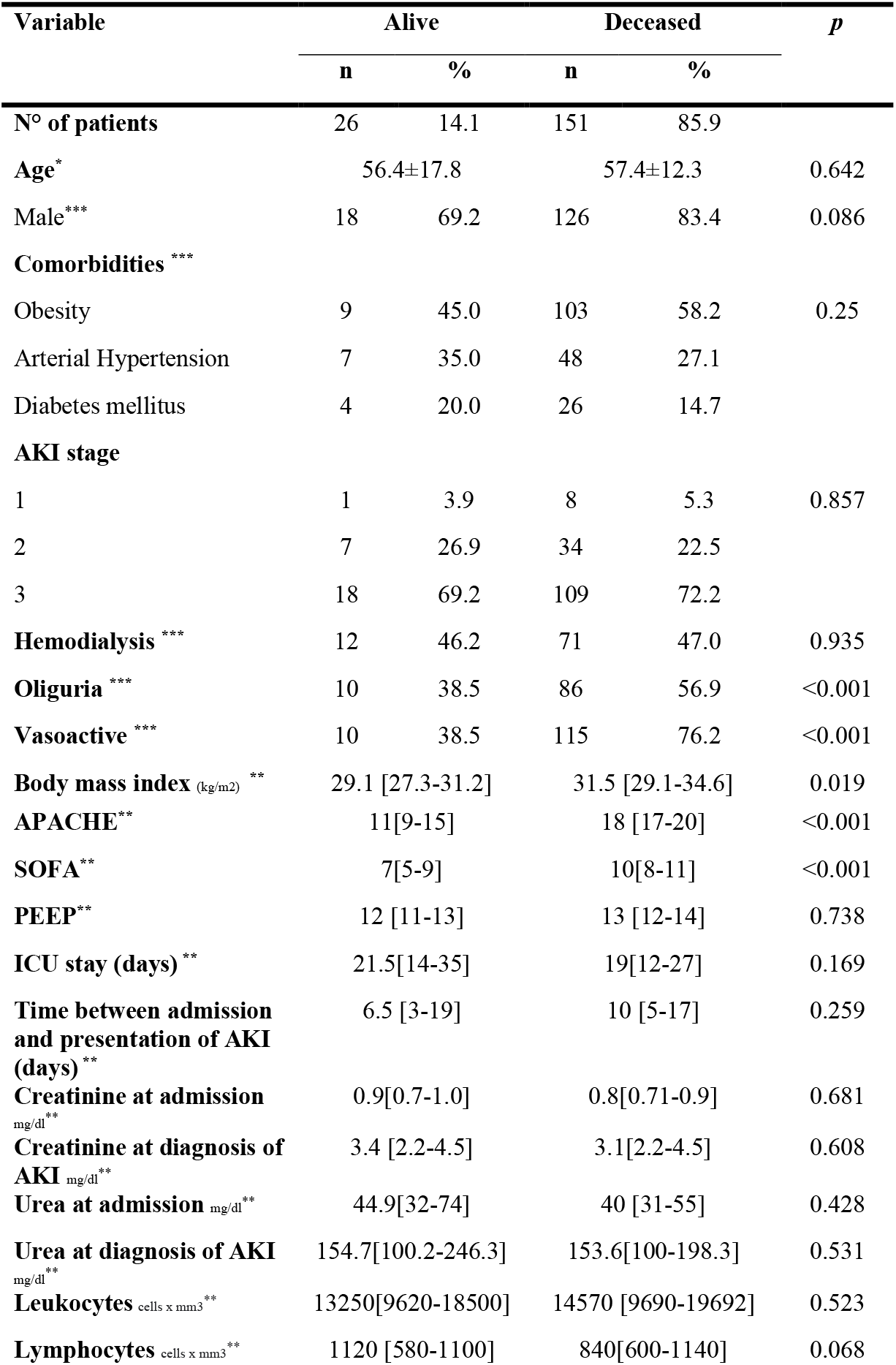
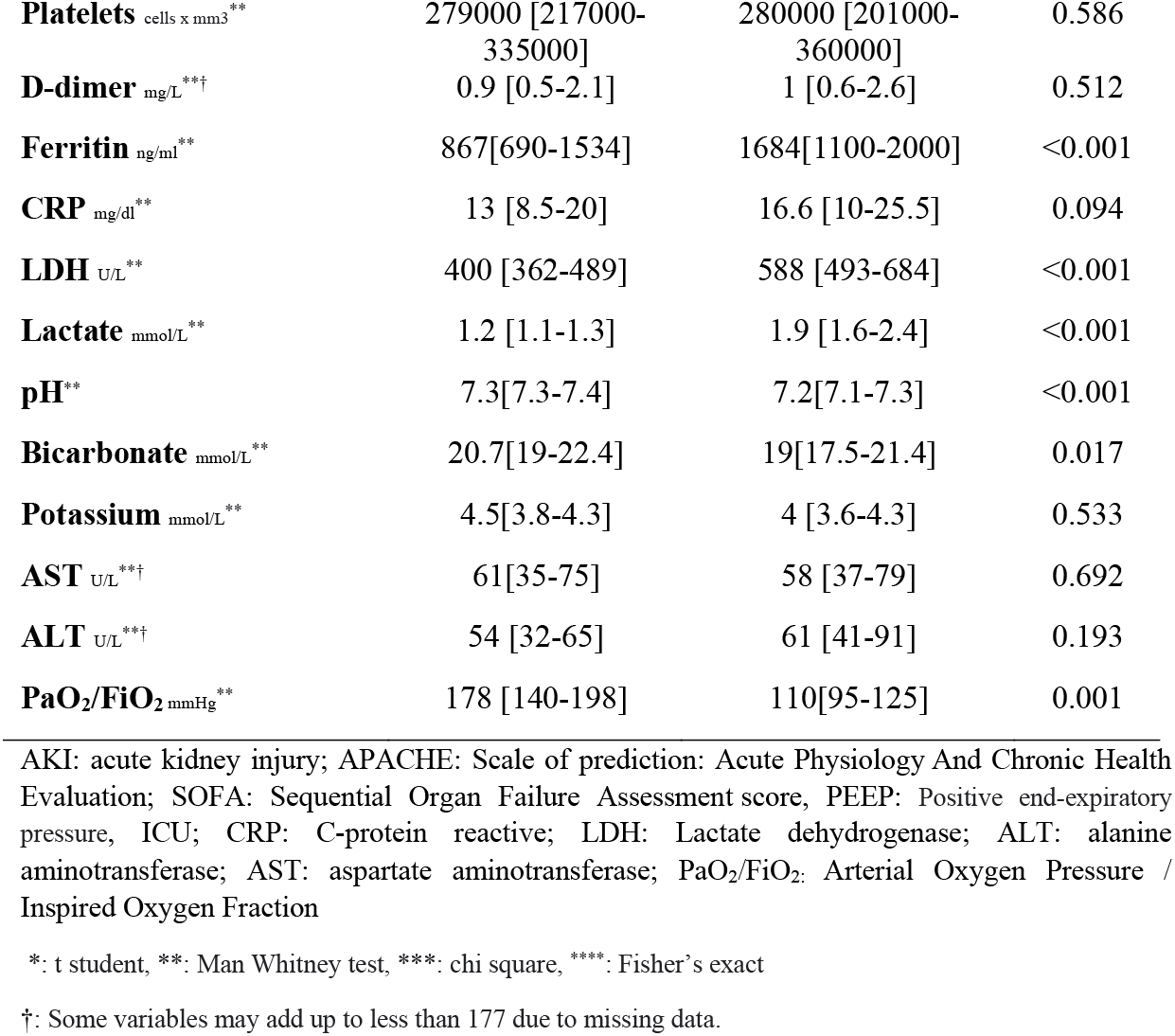
Clinical and laboratory characteristics, comparative between survivors and deceased.

In the exploratory multivariate analysis with logistic regression, increased ferritin level (OR: 10.04 (95%CI 4.4-38.46), p<0.001) and APACHE score (OR:1.75 (95%CI 1.4-2. 12), p<0.001), were related to mortality, while the higher PaO_2_/FiO_2_ level (OR: 0.85 (IC95% 0.59-0.92), p<0.042), was a protective factor; these results were adjusted for age, sex and comorbidities **(Figure 1)**.

**Figure 1.**
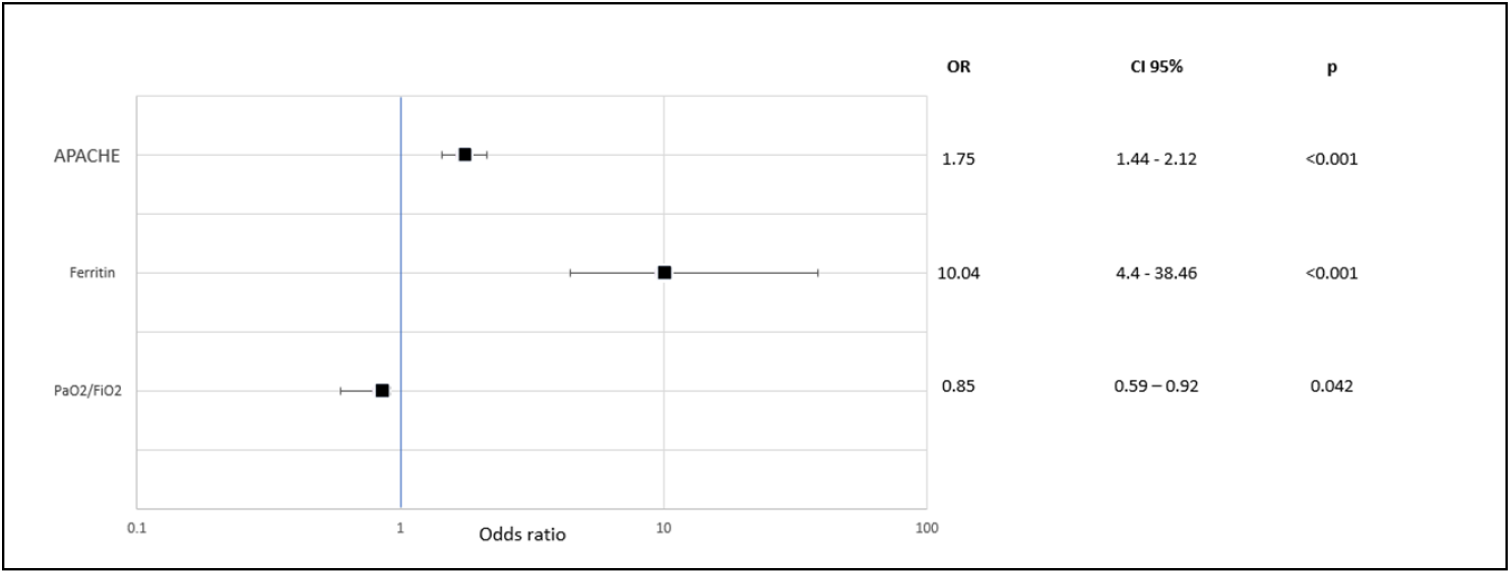
Factors related to mortality in patients with COVID-19 and AKI in ICU. APACHE: Scale of prediction: Acute Physiology and Chronic Health Evaluation. PaO_2_/FiO,_2_: Arterial Oxygen Pressure / Inspired Oxygen Fraction. OR: Odds ratio CI: Confidence interval Multiple logistic regression: adjusted for age, sex and comorbidities

## DISCUSSION

The present study evaluated 177 patients with COVID-19 infection hospitalized in the ICU and with AKI criteria, 46.9% received conventional hemodialysis and mortality was 85.3%.

The reported incidence of AKI by COVID-19 has varied over time, with initial reports mentioning frequencies of 3 to 9% (20), later reports have shown a higher frequency, both in hospitalized patients 36% to 46% (4, 21), and in those admitted to the ICU 40% to 69.8% (22-25). These differences may be explained by the characteristics of the populations analyzed and the definitions used for the diagnosis of ARF. This study only included patients with COVID-19 infection and who presented ARF in ICU and is comparable to two previous publications, with similar results in predominance of male patients (81.9% in our series), population of older adults between 57 and 71 years of age (26, 27) and not different from most published reports (23, 25).

Mortality was very high 85.3%, similar to that reported in similar populations (27,28), 4 important characteristics were identified in this group: a) the severity of renal compromise, manifested by oliguria which was present in 56.9% of the deceased, greater frequency of patients with ARF in stage 3, which was 71%, in contrast to the 17% to 51% reported in other studies (10, 23, 26) and the need for renal replacement therapy which was 46.9%. The presence of AKI in patients with COVID-19 is related to greater severity of the disease and higher mortality, with an OR of 8.45 and 13.5 respectively (29, 30); the need for dialysis support has been associated with a lower rate of recovery of renal function (31).

b) The severity of COVID-19 is another factor associated with mortality; patients who died had a greater need for vasoactive agents, a higher APACHE II score and SOFA, higher inflammatory markers ferritin (17), LDH and lactate, as well as a decrease in lymphocytes, of these factors ferritin and APACHE score were associated with mortality in multivariate analysis, similar to that reported by Taylor in a meta-analysis of 58 studies conducted in ICU, in which they refer to higher SOFA and APACHE-2 scores associated with mortality (32).

c) Pulmonary compromise is one of the most frequent and serious complications of COVID-19, the virus infects the alveolar cells through the receptors of the angiotensin converting enzyme 2 producing pneumonia and inducing severe immune response. In this study 100% of the patients required mechanical ventilation, higher than the 84 and 90% reported in similar studies (26, 27), the use of mechanical ventilation is associated with a mortality of 80% in hospital population (33), pronation is a maneuver that has been used 100% in patients with severe pulmonary compromise to improve oxygenation and was used in 100% of the patients in the study, and was higher than the 70% reported in other studies (34), PaO_2_/FiO_2_ levels were 113, values much lower than those reported (26) and was associated with high mortality in the multivariate analysis.

d) Comorbidities such as diabetes, arterial hypertension, chronic renal disease, obesity are associated with higher mortality in patients with COVID-19 (26,32,36,37), in our study the most frequent comorbidity was obesity with 63% (an average body mass index of 31 kg/m^2^), much higher than the 10 to 59 % reported in other studies, hypertension and diabetes mellitus was lower than reported 31% vs 45 to 64% (22, 25) and diabetes mellitus 16% vs 21 to 38% (26, 35) respectively.

The limitations of the study are: it is a retrospective study, whose primary source of data is the hospital clinical history, which implies some missing data; likewise, the group studied was small and not randomized, which could be improved with a better sampling strategy, observation time and the inclusion of other hospital centers, as well as the addition of all the patients admitted to ICU and not only those who presented ARF. The analysis of the factors related to mortality is only exploratory, it is not conclusive since we did not have an initial hypothesis, however, it is an opportunity to carry out other studies with a larger population and even follow-up of the survivors.

In conclusion, acute kidney injury in ICU patients with COVID-19 infection has a high mortality and the related factors were the increase in APACHE score and ferritin level, as well as the decrease in PaO_2_/FiO_2_ level.

## Supporting information

ETHICS

## Data Availability

Todos los datos producidos en el presente trabajo estan contenidos en el manuscrito.

## DISCLOSURE

All the authors declared no competing interests.

## FUNDING

Self-financing

## Notes

### Competing Interest Statement

The authors have declared no competing interest.

### Funding Statement

Este estudio no recibio ningun tipo de financiacion

### Author Declarations

The Institutional Research Ethics Committee of the Hospital Nacional Arzobispo Loayza approved the study under number 038-2021 and since it is a retrospective study, it is exempt from informed consent.

