## Supplementary material for "Acute renal injury in patients with COVID-19, in the critical care unit of a public hospital, Lima-Peru": ETHICS

"Decade of Equal Opportunities for Women and Men".
"Year of Unity, Peace and Development".

Lima October 18, 2021

### CONSTANCE 038-2021

The president of the Institutional Research Ethics Committee of the Hospital Nacional Arzobispo Loayza, hereby certifies that the research project indicated below was APPROVED by the CIEI under the category of EXPEDITED review.

Title of the research project: “Acute renal injury in patients with COVID-19, in the critical care unit of a public hospital, Lima-Peru”.

Principal Investigator(s): Joanna Yanissa Venegas Justiniano

The approval considers compliance with good clinical practices, current guidelines on ethics and scientific research in the field of health, risk/benefit balance and data confidentiality, among others.

MINISTERIO DE SALUD
HOSPITAL NACIONAL "ARZOBISPO LOAYZA"
Dr. Eduardo Matos Prado
Presidente del Comité Institucional de Ética en Investigación

EMP/
Jessica P.
Cc. Archivo
